# Problems of Deploying CNN Transfer Learning to Detect COVID-19 from Chest X-rays

**DOI:** 10.1101/2020.05.12.20098954

**Authors:** Taban Majeed, Rasber Rashid, Dashti Ali, Aras Asaad

## Abstract

The Covid-19 first occurs in Wuhan, China in December 2019. After that the virus spread all around the world and at the time of writing this paper the total number of confirmed cases are above 4.7 million with over 315000 deaths. Machine learning algorithms built on radiography images can be used as a decision support mechanism to aid radiologists to speed up the diagnostic process. The aim of this work is to conduct a critical analysis to investigate the applicability of convolutional neural networks (CNNs) for the purpose of COVID-19 detection in chest X-ray images and highlight the issues of using CNN directly on the whole image. To achieve this task, we first use 12-off-the-shelf CNN architectures in transfer learning mode on 3 publicly available chest X-ray databases together with proposing a shallow CNN architecture in which we train it from scratch. Chest X-ray images fed into CNN models without any preprocessing to follow the many of researches using chest X-rays in this manner. Next, a qualitative investigation performed to inspect the decisions made by CNNs using a technique known as class activation maps (CAM). Using CAMs, one can map the activations contributed most to the decision of CNNs back to the original image to visualize the most discriminating regions on the input image.

We conclude that CNN decisions should not be taken into consideration, despite their high classification accuracy, until clinicians can visually inspect, and approve, the region(s) of the input image used by CNNs that lead to its prediction.

## I. INTRODUCTION

The severe acute respiratory syndrome coronavirus 2 (SARS-CoV-2), the virus causing COVID-19, has become a pandemic since its emergence in Wuhan, China in Dec 2019 [1]. The death toll from the infection is escalating at a worrying rate and many health systems around the world are struggling to cope. Social distancing is among some approaches proposed by the World Health Organization (WHO) to control the spread of this viral infection. A critical step in this direction is an effective and accurate screening of the COVID-19 patients so positive cases receive timely treatment and get appropriately isolated from the public; a measure deemed crucial in curbing the spread of the infection. Reverse-transcription polymerase chain reaction (RT-PCR) testing, which can detect SARS CoV-2 RNA from respiratory specimens (such as nasopharyngeal or oropharyngeal swabs), is the golden screening method for detecting COVID-19 cases. The high sensitivity of RT-PCR testing is overshadowed by the limited availability of test kits and the amount of time required for the result to be available (few hours to a day or two) [2]. Therefore, there is a growing need to use fast and reliable screening techniques that could be further confirmed by the RT-PCR testing. Some studies have suggested the use of imaging techniques such as X-rays Computed Tomography (CT) scans of the chest to look for visual indicators associated with SARS-CoV-2 viral infection. It was found in early studies that patients display abnormalities in chest radiographs that are characteristic of COVID-19 infection, with some suggesting that radiography examination could be used as a primary tool for COVID-19 screening in epidemic areas [3]. Facilities for chest imaging is readily available in modern healthcare systems making radiography examination a good complement to RT-PCR testing and, in some cases, showing even a higher sensitivity index. Given X-ray visual indicators could be subtle; radiologist will face a great challenge in being able to detect those subtle changes and interpreting them accurately. As such, it becomes highly desired and required to have computer-aided diagnostic systems that can aid radiologists in making a more time-efficient and accurate interpretation of X-ray changes that are characteristic of COVID-19 [4].

In recent months, many researches came out addressing the problem of COVID-19 detection in chest X-rays using deep learning approaches in general, and convolutional neural networks (CNNs) in particular [3]–[10]. Majority of the papers report high COVID-19 disease detection accuracy [2][6][10]–[14]. For a detailed survey of recent artificial intelligence algorithms, we direct interested reader to check the well-written survey in [15] by Thanh Thi Nguyen. However, deploying CNN architectures directly on chest radiography images may not produce reliable COVID-19 detection results, especially when chest X-ray images feed into CNN models directly without any preprocessing steps such as region of interest segmentation, noise elimination and un-wanted object removal. We take this hypothesis onboard to prove that despite high classification accuracies by CNN models, we demonstrate that CNNs are ‘cheating’ by using artefacts in the images to build their prediction that has nothing to do with COVID-19 disease.

## II. Related Works (Survey)

Since the start of COVID-19, researchers quickly divided their effort on combating it by focusing on developing a vaccine in one hand [16] and detecting COVID-19 using PCR and imaging systems on the other hand [3]. Here, we review studies devoted to the use of radiography images to aid and complement PCR in diagnosing COVID-19 cases. Ai et al. [3] built a deep convolutional neural network (CNN) based on ResNet50, InceptionV3 and Inception-ResNetV2 models for the classification of COVID-19 Chest X-ray images to normal and COVID-19 classes. They reported a good correlation between CT image results and PCR approach. Chest X-ray images of 50 COVID-19 patients have been obtained from the open source GitHub repository shared by (Dr. Joseph Cohen [17]). Prabira et al. in [5] proposed a method to detect COVID-19 using X-ray images based on deep feature and support vector machines (SVM). They collected X-ray images from GitHub, Kaggle and Open-I repository. They extracted the deep feature maps of a number of CNN models and conclude that ResNet50 is performing better despite the small number of images used in their investigation. Maghdid et al. [6] proposed a simple CNN of 16 layers only to detect COVID-19 using both X-ray and CT scans and reported good performance but the dataset used is small. The work of Fei et al. [18] focused on segmenting COVID-19 CT scans using a deep learning approach known as VB-Net and reported dice similarity of 91%±10%.

Xiaowei et al. [8], obtained an early prediction model to classify COVID-19 pneumonia from Influenza-A viral pneumonia and healthy cases using pulmonary CT images using Resnet18 model by feeding image patches focused on regions of interest. The highest accuracy for the CNN model was 86.7 % CT images. In Wang et al. [9], authors use CT images to predict COVID-19 cases where they deployed Inception transfer-learning model to establish an accuracy of 89.5% with specificity of 88.0% and sensitivity of 87.0%. In [4] a number of CNN architectures that are already used for other medical image classifications evaluated over a dataset of X-ray images to distinguish the coronavirus cases from pneumonia and normal cases. CNN’s adopted on a dataset of 224 images of COVID-19, 700 of non- COVID19 pneumonia, and 504 normal where they report overall accuracy of 97.82.

Wang and Wong [2] investigated a dataset that they called COVIDx and a neural network architecture called COVID-Net designed for the detection of COVID-19 cases from an open source chest X-ray radiography images. The dataset consists of chest radiography images belonging to 4 classes including Normal X-rays comprising cases without any infections, Bacterial, Viral pertaining to non-COVID-19 pneumonia and COVID-19 X-rays. They reported an overall accuracy of 83.5% for these four classes. Their lowest reported positive predictive value was for non-COVID-19 class (67.0%) and highest was for Normal class (95.1%). As required to improve the previous studies Muhammad and Hafeez [7] deals with this need by presenting another CNN with fewer parameters but better performance. Authors used the same dataset as in [2] to build an open source and accurate COVID-ResNet for differentiating COVID-19 cases from the other four pneumonia cases and outperform COVID-Net. In [10], Narin et al. experimented several CNN architectures classify normal with COVID-19 X-ray images and they report excellent classification accuracy, sensitivity and specificity. But the authors failed to discuss the clinical importance of their approach as it may not be difficult to distinguish severe COVID-19 cases from normal chest X-rays, as we show in table 1, and this is not the situation radiologists face in a regular basis or it may not be of importance in this current pandemic. Finally, they trained their CNNs based on 50 images from each of the normal and COVID-19 classes which may result in some sort of biasness in the training phase.

**Table 1.** Testing result for all architectures used in scenario 1.

| CNN Architectures | Sensitivity | Specificity |
| --- | --- | --- |
| AlexNet | 90.41 | 88.03 |
| GoogleNet | 93.15 | 96.15 |
| Vgg16 | 84.93 | 97.86 |
| Vgg19 | 0 | 100 |
| ResNet18 | 95.89 | 98.72 |
| ResNet50 | 95.89 | 97.01 |
| ResNet101 | 91.78 | 97.86 |
| InceptionV3 | 97.26 | 92.74 |
| InceptionResNetv2 | 95.89 | 99.57 |
| SqueezeNet | 93.15 | 99.57 |
| Densenet201 | 90.41 | 100 |
| Xception | 93.15 | 100 |
| CNN-X (Ours) | 93.15 | 97.86 |

In all the works discussed here, to the best of our knowledge, we did not encounter an explicit description of preprocessing, segmentation nor noise reduction performed on chest X-rays. Also, whether such operations will affect the final decision by proposed CNNs or not. We address this problem by assessing the quality of the decision made by 12 CNN models using class activation mapping introduced in [19]. Also, there is no justification why researchers favored a particular CNN model over others and did not compare their final results if one opt to choose another CNN architecture. This paper benchmarks 12 popular CNN models and deploy them in a transfer learning mode on 3 public datasets popularized for the detection of COVID-19 infection. Finally, a qualitative analysis performed on these 12 CNN models to demonstrate the most discriminating regions in the input image used by each CNN and the need of such process to reveal the bias in current datasets as well as CNN weaknesses.

## III. CNN Architectures-Brief overview

In recent years, the use of deep learning algorithms in general and convolutional neural networks (CNNs) led to many breakthroughs in a variety of computer vision applications like segmentation, recognition and object detection [20]. Deep learning methods have been shown to be successful in automating the task of feature-representation learning and gradually attempts to eliminate the tedious task of handcrafted feature engineering. Deep learning, and convolutional neural networks (CNNs), attempts to mimic the human visual cortex system in terms of structure and operation by adopting a hierarchical layer of feature representation. This approach of multi-layer feature representation made it possible to learn different image features automatically and hence enabled CNNs to outperform handcrafted-feature methods [21].

In 1960s, Hubel and Wiesel [22] studied monkey’s visual cortex system and found cells which are responsible for constructing image and detecting light signal in receptive filed. In the same vein, Hubel and Wiesel also showed that monkey’s visual field can be represented using a topographic mapping. In 1980s, Neocognitron proposed by Fukushima and Miyake [23] which is a self-organizing neural network and regarded as a predecessor of CNN. In [24], LeCun et al.’s groundbreaking work introduced modern CNN models for the purpose of handwritten digit recognition in which the architecture later popularized and known as LeNet. After LeNet architecture, convolutional layers and backpropagation algorithm for training popularized and became a fundamental building block of most of the modern CNN architectures. In 2012, AlexNet architecture, proposed by Krizhevsky et al. [25], won ImageNet Large Scale Visual Recognition Challenge (ILSVRC) [26] by outperforming other methods and reducing the top-5 error from 26% to 15.3%. This was a turning point so that CNNs became an exceptionally popular tool to be deployed in many computer visions tasks. Roughly speaking, AlexNet is a similar version of LeNet but deeper structure and trained on 1.2 million high resolution images. Complex architectures that has millions of parameters, and hyperparameters, to train and fine tune need a substantial amount of computational time and power but again AlexNet popularized the use of powerful computational resources such as graphical processing units (GPUs) to compensate the increase in trainable parameters.

AlexNet opened the door for researchers around the world to design novel CNN models which are deep but efficient at the same time especially after ILSVRC became an annual venue for the recognition of new CNN models. The participation of technology giants such as Google, Microsoft and Facebook also helped in pushing research in this direction especially the depth of CNN architectures increased dramatically from 8 layers in 2012 to 152 layers in 2015 which helped the recognition error rate to drop to 3.5%. Pre-trained CNN architectures on ImageNet have been open-sourced and immediately used by researcher to transfer the knowledge to other application domains and promising results achieved [27]. One of the many useful features of transfer learning (TL) is that in other domains, such as medical image analysis, millions of labeled medical images are not available therefore it is natural to consider the use of fine-tuned weights and biases of CNN architectures trained on ImageNet, and other large databases, to be used for medical image analysis. Hence, we opt to use 12 deep learning architectures in a TL mode and modify their final layers to adapt to the number of classes in our investigation. The deep learning architectures that we used for the purpose of COVID19 detection from X-ray images are AlexNet, VGG16, VGG19, ResNet18, ResNet50, ResNet101, GoogleNet, InceptionV3, SqueezeNet, Inception-ReseNet-v2, Xception and DenseNet201.

In what follows we are going to briefly describe each of the 12 CNN architectures used here and highlight their distinct properties. It is out of the scope of this work to give details of all of these 12 CNN models, hence we direct interested reader to consult many survey articles on deep learning and CNN architectures such as [28], [29].

**AlexNet** architecture is the winner of ILSVRC 2012, proposed by Krizhevsky et al. [25] outperformed the handcrafted features significantly. AlexNet constitutes of 5 convolutional layers and 2 fully connected layers together with rectified linear unit (ReLU) activation function which is used for the first time. It can be regarded as a scaled version of LeNet except that it is a deeper architecture trained on a larger dataset of images (ImageNet) and benefitted from the GPU computational power. Hyperparameters of AlexNet fine-tuned and won 2013 ILSVRC [26] (later named ZF-Net). We use AlexNet in a transfer learning mode and modify the last layer of AlexNet according to the number of X-ray image classes, i.e. instead of 1000 classes that AlexNet trained on we change this to 4 classes because 4 X-ray classes used here which are COVID19, Bacteria, Viral and Normal. The same approach of TL is used for the rest of CNN models.

**VGG** architectures proposed by Oxford university’s visual geometry group [30], hence the acronym VGG, whereby they demonstrated that using small filters of size 3-by-3 in all of the convolutional layers throughout the network leads to a better performance. The main intuition behind VGG architectures is that multiple small filters in a sequence can imitate the effect of larger filters. Due to its simplicity in design and generalization power, VGG architectures are widely used. We use VGG16 and VGG19 that constitute of 16 and 19 convolutional layers, respectively.

**GoogleNet** architecture is the winner of ILSVRC 2014 which is proposed by Szegedy et al. [31] from Google in 2014. Novelty of GoogleNet is the innovation of inception module, which is a small network inside a bigger network. Furthermore, 1-by-1 convolutional layers/blocks used as a dimensionality reduction and feature aggregation. In total, GoogleNet is 22 layers deep with 9 inception modules. Inception V1 (GoogleNet), is later improved in terms of batch normalization, representational bottleneck and computational complexity and resulted in Inception V2 and V3. Here we opt to use GoogleNet and InceptionV3 [32] in a transfer learning mode. In the same vein, we use Xception [33], which is another architecture proposed by F. Chollet from Google which uses the idea of extreme inception module whereby depthwise convolutional layers used first then followed by pointwise convolutional layers. In other words, they replaced inception modules by depthwise separable convolutions in such a way that the total number of parameters is the same as inceptionV3 but the performance on large datasets (350 million images of 17000 classes) are significantly higher.

**ResNet** architectures are proposed by He et al. [34] from Microsoft and won 2015 ILSVRC. Main innovation in ResNet architectures are the use of residual layers and skip connections to solve the problem of vanishing gradient that may result in stopping the weights in the network to further update/change. This is particularly a problem in deep networks because the value of gradient can vanish, i.e. shrink to zero, when several chain rules applied consecutively. Skipping connections will help gradians to flow backwards directly from end layers to initial layer filters enabling CNN models to deepen with 152 layers.

**DenseNet** can be regarded as a logical extension of ResNet which was first proposed in 2016 by Huang et al. from Facebook [35]. In DenseNet, each layer of CNN connected to every other layer in the network in a feed-forward manner which helps in reducing the risk of gradient-vanishing, fewer parameters to train, feature-map reuse and each layer takes all preceding layer features as inputs. The authors also point out that when datasets used without augmentation, DenseNet is less prone to overfitting. There are a number of DenseNet architectures, but we opt to use DenseNet201 for our analysis of COVID19 detection from X-ray images by using the weights trained on ImageNet dataset in TL mode.

**SqueezeNet** is a small architecture proposed by Landola et al. [36] in 2016 that uses the idea of fire module which contain 3 filters of size 1-by-1 feed into an expanded layer (4 filters of size 1-by-1 and 4 filters of size 3-by-3). Even though the number of parameters of SqueezeNet is by 50x less than AlexNet but achieves the same accuracy of AlexNet on ImageNet.

**Inception-ResNetV2** is a combined architecture proposed by Szegedy et al. [32] in 2016 that uses the idea of inception blocks and residual layers together. The aim of using residual connections is to avoid the problem of degradation causes by deep networks and reduce the training time. The inception-resnetV2 architecture used here contains 20 inception-resnet blocks that empower the network to become 164 layers deep, and we use the pre-trained weights in these layers to assist our mission of detecting COVID19 in X-Ray images.

## IV. Proposed CNN

In this study, we designed a CNN model for COVID-19 detection from chest radiography images guided by the fact that in order to properly classify and detect COVID-19, radiologists need to discriminate COVID-19 X-rays from normal chest X-ray first, and then from other viral and bacterial infections in order to isolate and treat the patient properly. Therefore, we opt to choose the design of CNN to make one of the following predictions: a) Normal (i.e. no infection) b) COVID-19, c) Viral infection (none-COVID-19) and d) Bacterial infection. The rationale behind using these 4 cases is to aid radiologists to prioritize COVID-19 patients for PCR testing and employ treatments according to infection-specific causes. Having these requirements in mind, we designed our simple CNN architecture, named CNN-X, that constitutes of 4 parallel layers where we have 16 filters in each layer in 3 different sizes (3-by-3, 5-by-5 and 9-by-9). Batch normalization and rectified linear unit (ReLU) is then applied to the convolved images and two different types of pooling operation applied next which are average pooling and maximum pooling. The rationale behind using different filter sizes is to detect local-features using filters of size 3-by-3 and rather global features by filters of size 9-by-9 while 5-by-5 filter size is to detect what is missed by the other two filters.

Different pooling operations utilized to further reduce the dimensionality of feature maps. A stride of size 3 is adopted here, with pooling operations, to further reduce the dimension of the resulting feature maps taking into consideration the fact that there is redundant information in images and neglecting a row and a column after each pooling window is not causing a massive information loss. See Fig. 1 where we visually depict the difference between pooling of size 3-by-3 with stride 2 versus pooling of size 2-by-2 with stride 3 and conclude that we are not losing much information while reducing the size of the image/feature map further. Proposed architecture design is not deep, hence the feature map (i.e. convolved image) is not a very abstract representation of the input image yet and as such there are still redundant information.

**Fig. 1.**
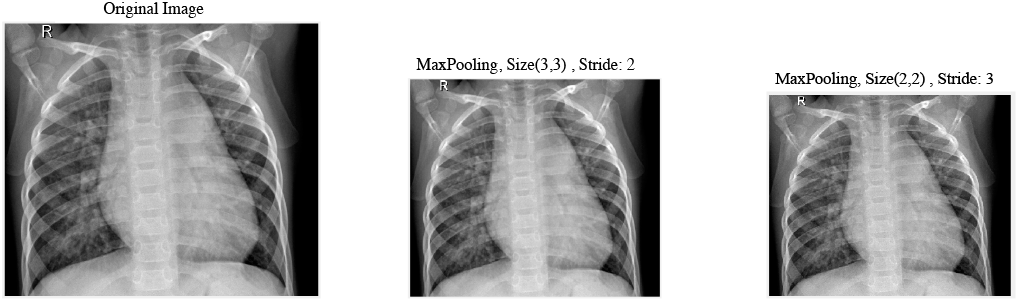
Effect of stride and pooling on image resolution.

Feature maps from the four parallel layers are then concatenated before fully connected layer. Weights are generated using Glorot method [37] with Adam optimizer [38] and 0.0003 initial learning rate. Training conducted using 20 epochs and 15 mini batch size. We visualize the structure of proposed CNN model in Fig. 2.

**Fig. 2.**
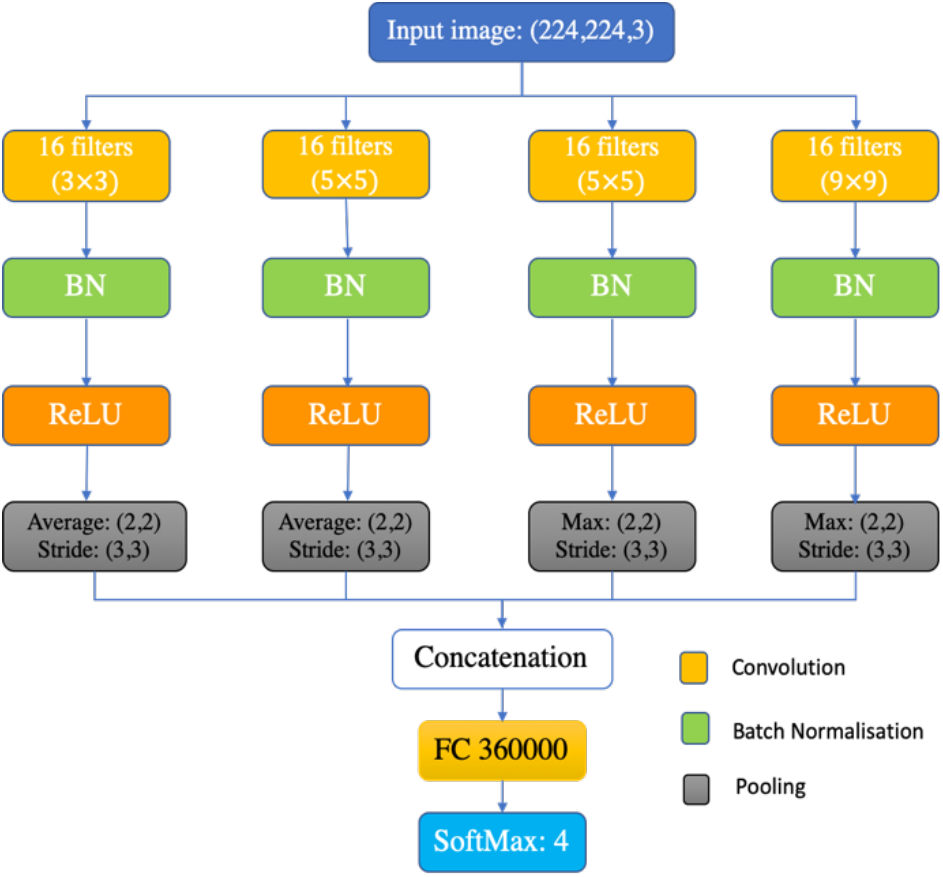
Proposed CNN Architecture design.

## V. Dataset Description

To investigate and test the CNN architectures explained in section III and IV, we used X-ray images collected from 3 publicly available sources. First dataset is a collection of 111 COVID-19 chest X-ray images collected by Cohen [17]. Second dataset is a collection of 6290 chest X-ray images of confirmed normal, bacterial and other non-COVID-19 viral infections from Kermany et al. [39]. Specifically, the images divided into four classes as follows; the total number of normal cases are 1575 cases, confirmed bacterial infection cases are 2771 and viral (Non-COVID-19) are 1494 confirmed cases. The third dataset contains 73 confirmed COVID-19 chest X-rays collected from the following websites; Radiological Society of North America (RSNA), Radiopedia, and Italian Society of Medical and Interventional Radiology (SIRM). This dataset again is available publicly in [40]. In Fig. 3 examples of all four radiographic X-ray classes are shown.

**Fig. 3.**
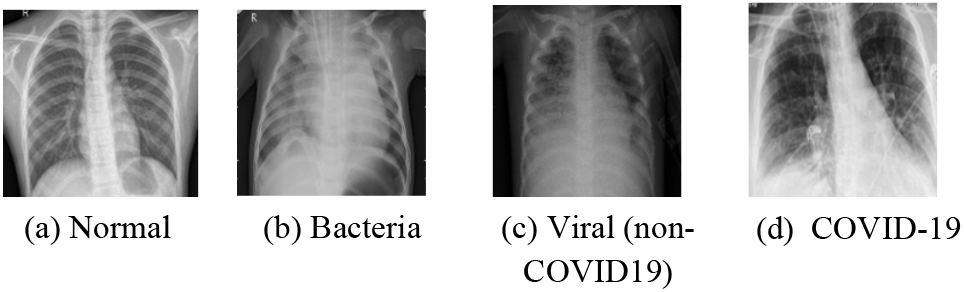
Sample of the X-Ray images used in our experiments.

Details of distributing the images to train set, validation set, and test set will be discussed and explained in the next section.

## VI. Experimental Setup and Results

We adopted transfer learning (TL) approach to investigate the performance of the CNN architectures discussed here and compare it with proposed CNN-X architecture. TL is the process of utilizing gained knowledge (learned weights) from solving one problem to a different but related problem. Weights optimized from training the 12 CNN models on ImageNet dataset used in TL mode such that weights in all layers are retrained on our X-ray images. All images from training and testing sets are resized to the suitable dimensions that each of the architectures designed for. No preprocessing applied to input images because none of the methods in literature work mentioned it and hence, we followed the same norm. Training parameters in TL for all 12 CNN architectures are as follows: number of epochs = 20, mini-batch size = 15, initial learning rate =0.0003. All experiments conducted using MATLAB version 2019b on a Core i5 CPU machine with 16 GB of RAM and 3.30 GHz. To measure CNN classification performance, four metrics were recorded which are sensitivity, specificity, F1-score and classification confidence. To be able to calculate the aforementioned metrics the following measures of test classification computed:

True positive (TP): number of correctly identified disease X-ray images.

False Negative (FN): number of incorrectly classified disease X-ray images.

True Negative (TN): number of correctly identified healthy X-ray cases

False Positive (FP): incorrectly identified healthy X-ray cases.

Furthermore, TP refers to disease (COVID-19, bacterial or viral) X-ray images correctly identified as a disease X-ray image while FP is normal or other pneumonia cases incorrectly identified as COVID-19 disease. Sensitivity measures the proportion of diseased cases correctly detected by CNNs while specificity measure the proportion of healthy cases correctly identified as healthy by CNN models. The equation of sensitivity and specificity calculation is provided in appendix, which also contain the F1-score calculation and equation. Because the number of COVID-19 chest X-ray images is small in comparison with the other 3 classes, it is sometimes misleading to rely on sensitivity and specificity of CNN models alone. Therefore, we also report the computation of the estimate of 95% confidence interval (see the appendix) of classification errors of each of the CNN models utilised here where we assume that the CNN classification output distributed normally, i.e. follows a gaussian distribution. The smaller the confidence interval, more reliable the predictive model is and hence one expects its CNN model more likely to work on other datasets.

Three different scenarios deployed to test the performance of 12 off-the-shelf CNN architectures as well as our proposed CNN-X model which will be discussed next.

### • Scenario 1: Normal vs COVID-19 classification (All Data)

In this scheme, CNN architectures trained on 1341 normal X-ray images with 111 COVID-19 cases while 234 cases of normal with 73 cases of COVID-19 are used for testing. Table 1 below shows obtained results from all the 13 CNN architectures. The aim of testing this hypothesis is to see the effect of differentiating COVID-19 from normal chest X-rays.

It can be seen from the table above that all of the CNN models (except Vgg19 and Vgg19), can be deployed successfully to detect COVID-19 X-rays with sensitivity of above 90%. However, the specificity of some of the techniques are below 90% in which we can avoid using it in practice. In this vein, one can opt to rely on the highest performing architectures such as Xception, Desnsenet201, SqueezeNet and inceptionresnetv2 as their specificity is >99%. It should be noted that our proposed CNN architecture’s performing is comparable to other state-of-the-art CNN models whereby it achieves 93% sensitivity and specificity of 97%, which is better than AlexNet, GoogleNet, VGG19 and VGG16. Albeit excellent results in table 1, this is not a realistic scenario to build machine learning algorithms for the purpose of COVID-19 detection in the present time because there is no guarantee that the system is not classifying other pneumonia infections as COVID-19 and vice versa. Furthermore, it may not be of a clinical significance to differentiate extreme COVID-19 cases from normal chest X-rays but it’s the diagnostics and discrimination of COVID-19 from other pneumonia is of a particular interest. Hence, we designed the second scenario to address the task of discriminating COVID-19 cases from other viral, bacterial and normal X-rays images.

### • Scenario 2: Normal vs COVID-19 vs Viral (non-COVID-19) vs Bacteria

In this scenario we aim to classify X-ray images into the 4 respective classes of normal, COVID-19, Bacteria and Viral (non-COVID-19). This scenario addresses the limitation in the first scenario whereby any machine learning algorithm needs to, ultimately, discriminate not only COVID-19 chest X-ray from normal X-ray but it also needs to discriminate COVID-19 chest X-rays from other viral and bacterial infections. This is a necessary condition to stop the spread of the virus and prepare COVID-19 patients for special treatments.

A total of 1341 normal X-rays, 2529 Bacteria cases, 1346 Viral X-rays and 111 COVID-19 X-rays used for training. For testing, 234, 242, 148 and 73 X-rays of normal, Bacteria, Viral and COVID-19 respectively used. It is worth to notice that we train the model on 111 COVID chest X-rays from COVIDx dataset but we test the CNN models on 73 chest X-rays from a different source. This is critical to examine the effectiveness of feature maps learnt by CNN on one source and testing it on images coming from a different source. Table 2 below demonstrates classification performance obtained by adopting this scenario.

### • Scenario 3: Normal vs COVID-19 vs Viral vs Bacteria (Training on part of the data)

In this scenario we used part of the dataset to train CNN models to see the effect of each architecture with the smaller number of image samples. The rationale behind this scenario is the fact that most of the time the challenge in medical image analysis is limitation of available data for investigation and to reduce bias in having unbalanced number of images in training phase. Hence, the design of this scenario is to get more insight of how these CNN models perform in the case of limited availability of image samples.

In this scenario, four classes used with 350 X-ray images of normal, Bacteria, viral and 111 X-rays of COVID-19 for training whereas the same number of testing images used for the four classes are as scenario 2.

Table 2 shows experimental results obtained from scenario 2 and scenario 3, where S_n_ and S_p_ stand for sensitivity and specificity respectively in Table 2. It clearly depicts that none of the CNN architectures perform well on differentiating X-rays to all four classes. Perhaps the only exception is Inception-ResnetV2 that performs better in comparison with the rest of the architectures especially on normal X-rays with sensitivity of >76% using all image samples. The good performance of Inception-ResnetV2 is due to the idea of combining residual learning with inception blocks which makes the performance to be better than ResNet or Google/Inception architectures alone. Furthermore, we notice that all CNN models work well on detecting two of the classes, namely Bacteria and COVID-19, but not performing well on classifying normal and viral X-rays to their respective classes which suggests that deployed CNN models learns features of bacterial and COVID-19 better than normal and non-COVID19 viral infections.

**Table 2.**
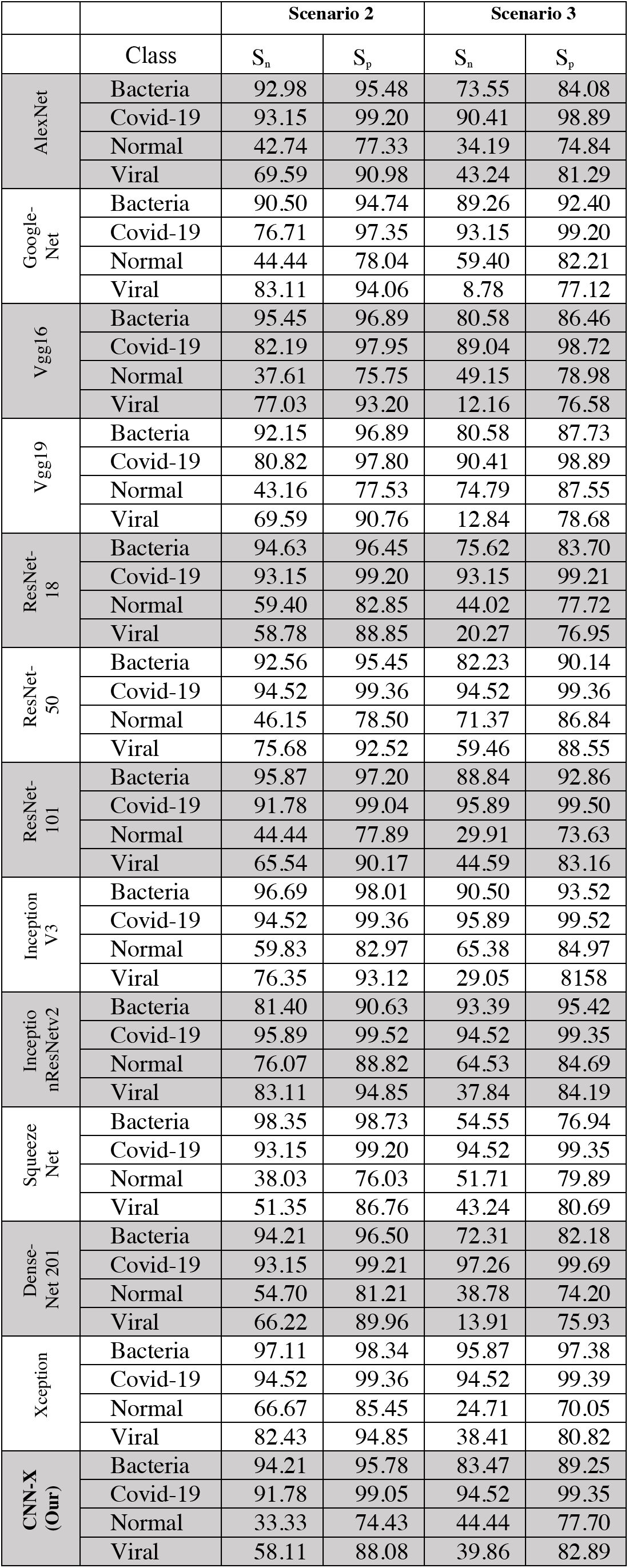
Testing result for scenario 2 and scenario 3 for all Models.

In other words, there is more similarity between features of X-ray images of viral infection and normal cases with each other and with other classes that cannot be distinguished easily. The second-best performing architecture, using all image samples, is Xception architecture with sensitivity of 97%, 94%, 66% and 82% for bacteria, COVID-19, normal and viral chest infections respectively. When it comes to scenario 3, where only 350 images used from normal, bacterial and viral chest X-rays, again Inception-ResnetV2 outperform all other CNN architectures including CNN-X. This confirms the effectiveness of Inception-ResnetV2 in terms of design and learning power. Nonetheless, we want to remind the reader that input images have not been segmented and they contain artefact that may contribute to CNN prediction but has no relation to COVID-19 infection. We confirm this point in the next section, see Fig. 4, where we demonstrate the region(s) in the image used by CNNs and some, if not all, of these regions are artifacts. Direct comparison of best results obtained here, which is by Inception-ResnetV2, is not possible with other works in the literature because the COVID-19 images used for testing here is different and more importantly the number of testing images is 73 which is higher than the number of test images used in [2] and [7] whereby they tested their CNNs based on 8 COVID-19 images only. Nonetheless, our results are outperforming COVID-Net in terms of sensitivity for viral and normal X-ray classification. The sensitivity of Inception-ResNet-V2 is again outperforms COVID-Net for bacterial, COVID-19, and viral infection classification.

Proposed CNN-X architecture is performing like Squeeze-Net, which is an architecture with 1.2 million parameters and our architecture has about 1.4 million parameters. Furthermore, CNN-X’s performance is comparable to VGG16 and AlexNet architecture which indicate that shallow CNN networks has the potential to be used to detect COVID-19 in the future with further improvements.

Next, we analyse qualitatively the performance of all CNN models used to visually inspect the most discriminating regions they utilized on classifying input chest X-rays into a specific category. This step is critical so that radiologists can visualize the regions used by CNNs to predict pneumonia presence in input X-ray images.

## VII. CNN interpretability

There are many ways one can visualize the region(s) used by CNNs to predict the class label of an input image such as gradient descent class activation mappings or global average pooling class activation mappings and others [19][41][42].To interpret the output decision made by any of the CNN architectures investigated in this study, heatmaps of the most discriminating regions generated and visualized for the input images in testing using the method introduced in [19] which is known as class activation mappings (CAM). Using CAMs, one can highlight class specific distinctive regions used by CNNs that lead to its prediction. After fully training a CNN model, a testing image will be feed into the network and feature maps extracted from final convolutional layer. In what follows we briefly introduce the steps of generating CAMs. Let *A_u_*(*x*, *y*) be activation of unit *u* of the last convolutional layer at a spatial position of (*x, y*). Let

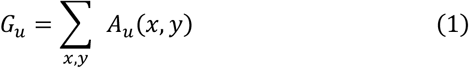

be average pooling operation and the input by the SoftMax layer is then can be defined as follows:

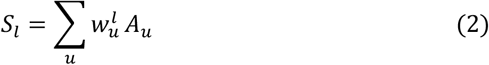

where *l* is the class label, 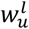 is the weight of class *l* of the unit *u*. Here, 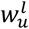 highlights important of the activation *A_u_* for a given class *l*. Probability score output by SoftMax for a given class *l* can then be defined as follows:

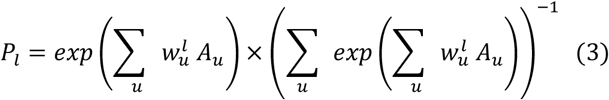

Substituting equation (1) into equation (2) we obtain the following:

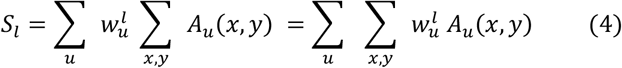

Then each class *l* activation maps can be defined at each spatial position (*x, y*) as follows:

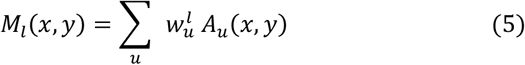

Finally, substituting activation maps for each class label in equation (5) into equation (4) we obtain the activation output by SoftMax for each class label *l* as follows:

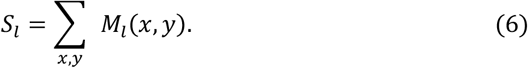

Hence, *M_l_*(*x,y*) indicates the discriminative power of activation maps at the spatial grid (*x,y*) that leads to the decision made the CNN to classify the input image into class *l*. To allow comparison to the input image, bilinear up-sampling is then applied to resize activation map to the size of input images accepted by each CNN model.

In Fig.4 we demonstrate the image regions used by CNN models that lead to a successfully class prediction. It can be observed that in very few occasions the CNN algorithms are focusing on the frontal region of the chest where we search for signs/features of COVID-19 and other infections. Rather, they are using either regions outside the frontal view of chest area, see row (d) and (e) of Fig. 3, or texts and medical device pieces on the images to derive their decision, see 1^st^ column in row (b) of Fig. 4.

In the same vein, incorrect classification, too, may be caused by these artifacts, see Fig 5 where we show examples of mis-classified images by CNNs and their corresponding CAMs to highlight the most discriminating regions lead to CNN decisions. For example, in 4-column of Fig. 5 is an X-ray image that has a letter *R* on it and used by almost all the architectures to drive their incorrect decision. Therefore, we conclude that using X-ray images as it is, without preprocessing to segment the region of interest and remove some hidden noise, is not a good practice and result in a biased and misleading classification prediction. In other words, one wants to have a CNN model that learn the symptoms of COVID-19 and its classification prediction is solely based on these features.

**Fig. 4.**
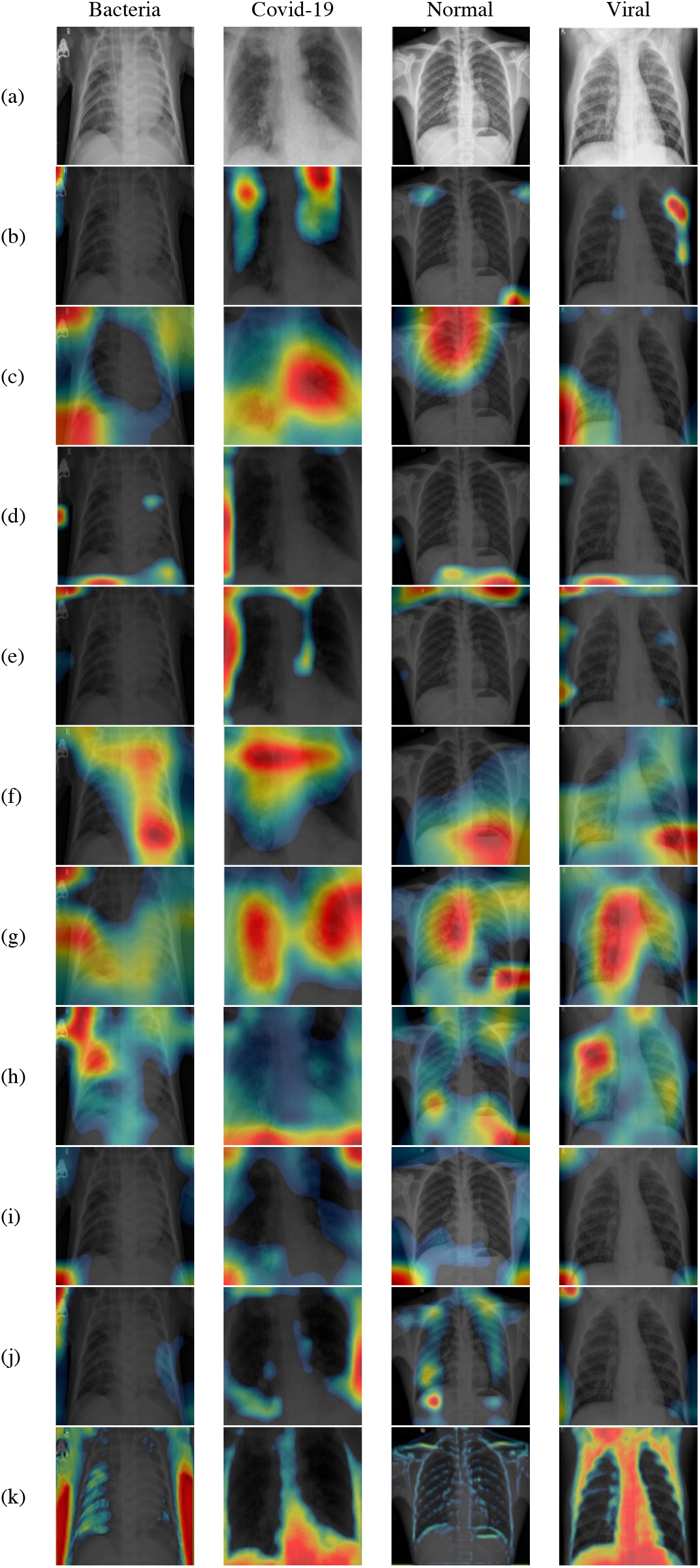
Visualization of X-rays images classified correctly by CNNs. (a) Original X-ray, (b) AlexNet, (c) GoogleNet, (d)VGG16, (e) VGG19 (f) ResNet18, (g) ResNet50 (h) InceptionResNet (i) DenseNet (j) SqueezeNet and (k) CNN-X (ours).

**Fig. 5.**
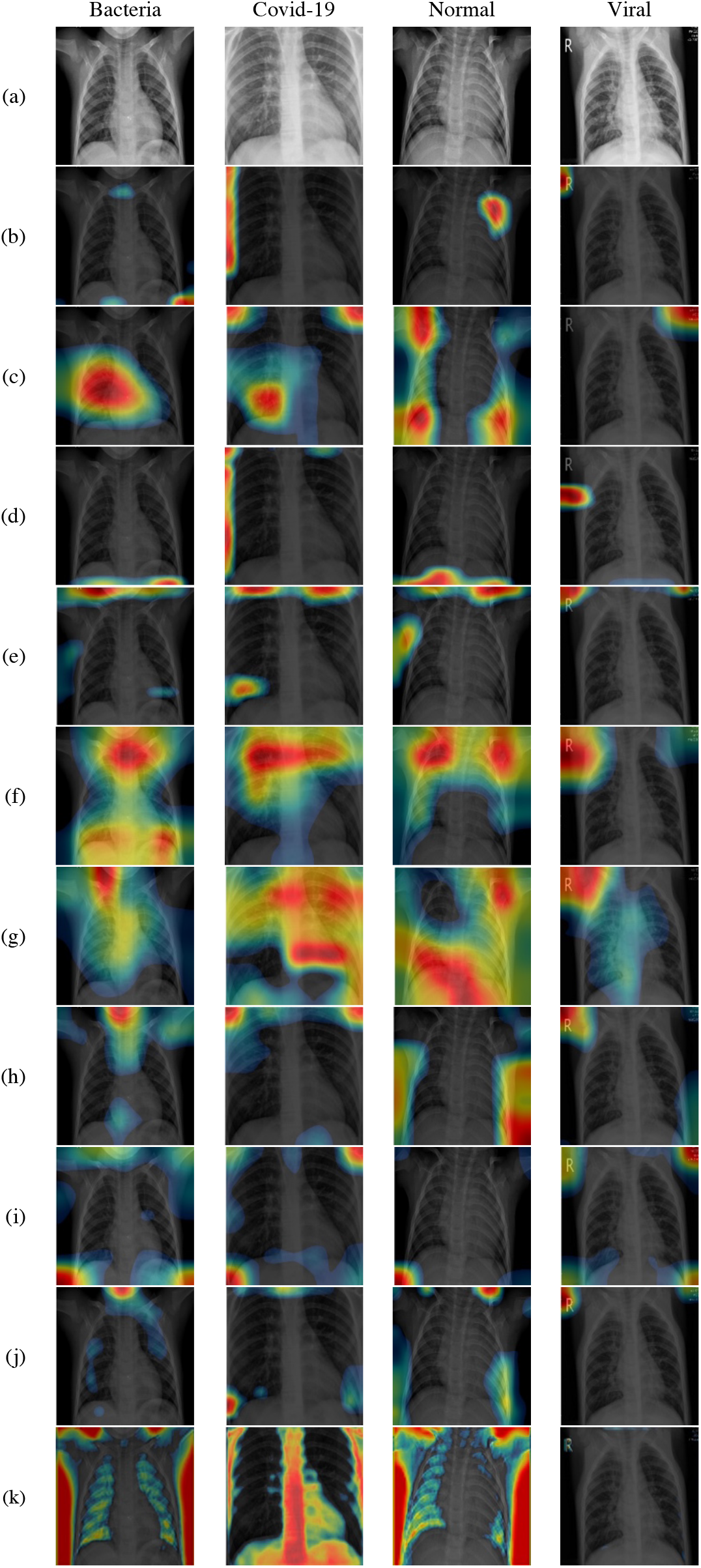
Visualization of X-rays images classified incorrectly by CNNs. (a) Original X-ray, (b) AlexNet, (c) GoogleNet, (d)VGG16, (e) VGG19 (f) ResNet18, (g) ResNet50 (h) InceptionResNet (i) DenseNet (j) SqueezeNet and (k) CNN-X (ours).

## VIII. Conclusion and Future Works

This paper presented a critical analysis for 12 off-the-shelf CNN architectures, proposed originally for natural image analysis, for the purpose of aiding radiologists to discriminate COVID-19 disease based on chest X-ray images. We also proposed a simple CNN architecture, with fewer parameters, that can outperform architectures such as Xception and Dense net when trained on a small dataset of images. Furthermore, beside quantitative analysis of CNNS, we qualitatively assessed CNN methods investigated in this paper using class activation mappings where we visualize the regions on X-ray images utilised by CNNs to derive their final prediction scores. We demonstrated that deep learning predictions of COVID-19 disease are not reliable when clear artefacts such as texts and medical device traces are present on the input X-ray image. In the same vein, we demonstrated that CNNs will use regions/features in the input image which are outside the ROI and have no relation with COVID-19 pneumonia, see Fig 5 for more than one example of this case as evidence.

Therefore, positive or negative class predictions by CNNs must be treated cautiously unless qualitatively inspected and approved by radiologists. Figures 4 and 5 contain multiple examples where *texts, medical device traces* and *irrelevant X-rays image regions* of *CODIVD-19 disease*used by CNNs to build their prediction result. It is important to note that, one needs to design machine learning algorithms based on radiologist opinion and not fully depend on data-driven mechanisms.

Future research directions, and in progress work, contain segmenting the lung region from chest X-rays and removing other artefact such as text and medical device traces on chest X-rays. We have not encountered any study that segmented the lung region in X-ray images and then feed it to CNN models, while this is considered as one of the important areas that needs to be further researched. The reliability of lung segmentation approaches is another problem that needs to be addressed and further researched by machine learning community. We have also not encountered, to the best of our knowledge, any study incorporated clinical and cardiac features with deep learning models or used cardiac features alone to prognosticate COVID-19 pneumonia. Data from other sources need to be incorporated to build CNN models that can be generalized and not biased towards a specific country, such as China or Italy, or a targeted population.

## Data Availability

All data and trained Models are available upon request.

https://github.com/ieee8023/covid-chestxray-dataset

## Appendix

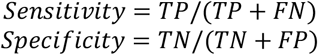

For Multi-Classes:

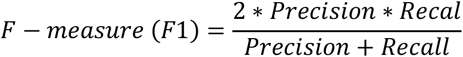

Where

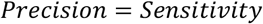

And

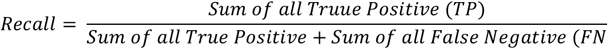

Confidence Interval of Classifier’s performance is equal to

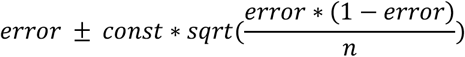

Where

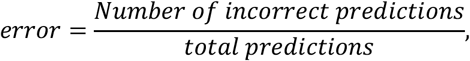

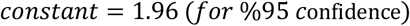

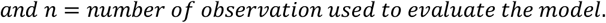

**Table 3.**
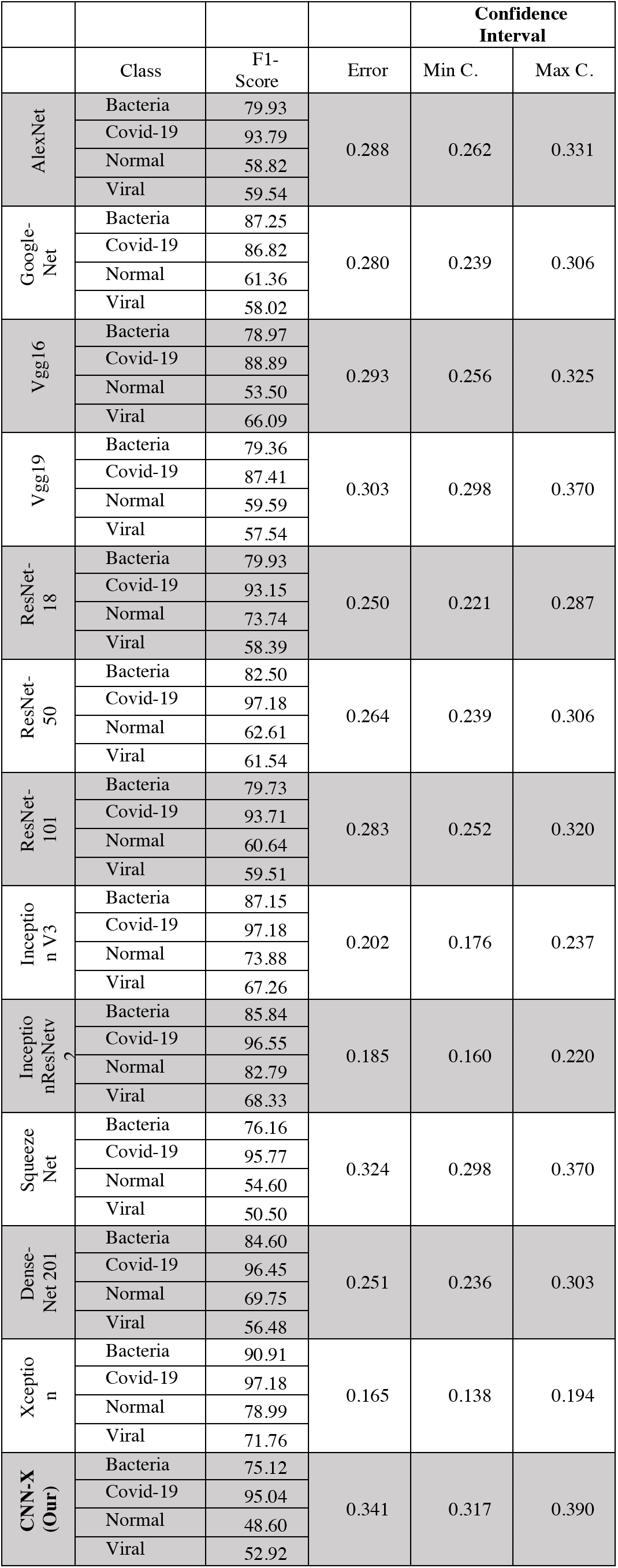
F1-Score and Classifiers confidence interval.

